# EFFECTIVENESS OF BASELINE AND POST-PROCESSED CHEST X-RAY IN NONEARLY COVID-19 PATIENTS

**DOI:** 10.1101/2020.04.16.20061044

**Authors:** Michele Gaeta, Giuseppe Cicero, Maria Adele Marino, Tommaso D’Angelo, Enricomaria Mormina, Silvio Mazziotti, Alfredo Blandino, Giulio Siracusano, Aurelio La Corte, Massimo Chiappini, Giovanni Finocchio

## Abstract

**Background:** CT is a very sensitive technique to detect pneumonia in COVID-19 patients. However, it is impaired by high costs, logistic issues and high risk of exposure.

Chest x-ray (CXR) is a low-cost, low-risk, not time consuming technique and is emerging as the recommended imaging modality to use in COVID-19 pandemic.

This technique, although less sensitive than CT-scan, can provide useful information about pulmonary involvement.

**Purpose:** To describe chest x-ray features of COVID-19 pneumonia and to evaluate the sensitivity of this technique in detecting pneumonia. A further scope is to assess the effectiveness of a post-processing algorithm in improving lung lesions detectability.

**Materials and Methods:** 72 patients with laboratory-confirmed COVID-19 underwent bedside chest X-ray.

Two radiologists were asked to express their opinion about: (i) presence of pneumonia (negative or positive); (ii) localization (unilateral or bilateral); (iii) topography (according to pulmonary fields); (iv) density (non consolidative ground-glass or inhomogeneous opacities; consolidative nodular-type or triangular; mixed consolidative e non-consolidative); and (v) presence of pleural effusion. The point (i) was evaluated separately, while the other points in consensus.

A quality assessment of post-processed x-ray images was performed by two different readers.

**Results:** The agreement about presence of pneumonia was almost perfect with K value of 0.933 and p < 0.001.

Sensitivity was 69%.

The following findings were seen: unilateral lung involvement in 50%; lower lung lesions in 54%; peripheral distribution in 48%; and non-consolidative pattern in 44%.

Post-processed images improved the detection of lesions in 7 out 72 patients (≅10%)

**Conclusion:** CXR owns a good sensitivity in detecting COVID-19 lung involvement. Use of post-processing algorithm can improve detection of lesions. Our data support recommendations of the Radiological Society of North America (RSNA) to consider chest x-ray as first step imaging examination in Covid-19 patients.

**SUMMARY:** Bedside CXR has a good sensitivity in evaluating COVID-19 lung involvement in hospitalized patients and should be considered as the first step imaging technique according to RSNA recommendations.

**KEY RESULTS:**

- Bedside CXR has a good sensitivity in evaluating COVID-19 lung involvement in non-early clinical cases.
- The most common findings of lung involvement were slight different from the well-described CT-ones, with less common patterns of bilateral and peripheral distribution.
- Post-processing algorithm enhances detection of pulmonary lesions.

## INTRODUCTION

The role of imaging in COVID-19 patients is rapidly evolving. Transcription-polymerase chain reaction (RT-PCR) which detects viral nucleic acid is the current and only reference standard in the diagnosis of COVID-19 according to World Health Organization [1].

Several papers have demonstrated which non-contrast chest computed tomography (CT) owns high sensitivity in detecting novel COVID-19 pneumonia, also in patients with initial false negative RT-PCR, suggesting the extensive use of chest CT to decrease false negative lab studies [2-9].

Although very sensitive in detecting pneumonia, CT is not specific for COVID and increases the risks for hospital community [10]. Moreover procedures of decontamination are difficult and time consuming and can disrupt CT availability for non-COVID19 patients.

For those reasons, since the beginning of the pandemic in Messina (Italy) (March 07th 2020), we have defined and started an imaging protocol for patients with COVID-19. It does not include routine CT but it is based on execution of chest x-ray (CXR) exclusively at the bed of patients after hospitalization in the COVID-center of the University Hospital of Messina.

The correctness of our approach has been confirmed by the North American Radiology Scientific Expert Panel which assessed that portable CXR has to be considered as the main imaging approach in evaluating COVID patients.

We have found only a paper from China describing CXR in a COVID-19 cohort of 64 patients [11]. We strongly believe that description of a further cohort of 72 patients studied with a CXR can be useful for radiologist involved in the diagnosis of COVID-19 lung involvement.

The aims of this paper are:

1. to describe CXR features of COVID-19 pneumonia in a cohort of 72 consecutive COVID-19 patients diagnosed with RT-PCR;
2. to evaluate the sensitivity of this technique in detecting pneumonia.

It is well known that a not negligible percentage of examinations performed in such a condition are not of perfect quality, mainly for the poor or absent collaboration of patients, thus a further scope of this work is related to introduce and assess the effectiveness of a state-of-the art post-processing algorithm for increasing the detectability of pulmonary lesions on CXR performed at bed.

## MATERIAL AND METHOD

We retrieved and analyzed CXRs obtained at the bed of 72 patients admitted in our COVID-center (25 females, 47 males, age range 43-100 years, mean age 68.8 years). Each patient resulted positive for COVID-19 by RT-PCR testing (in one case biological material was obtained by bronchioloalveolar lavage –BAL).

Clinically, the patients have been divided in 3 groups hospitalized in 3 different areas of our hospital according to criteria for admission in our COVID-19-center:

1. almost severe with fever, cough, slight shortness of breath at rest or under exertion (34 patients)
2. severe with reduced blood oxygen saturation (not inferior 90%) requiring oxygen therapy by mask (29 patients)
3. critical with respiratory failure requiring intubation and treatment in intensive therapy care unit (9 patients)

No patients with mild symptomatology was admitted because they were kept in quarantine and treated at home unless a worsening of their symptomatology due to biphasic evolution of the disease.

All the patients underwent examination within 24 hours from admission in the center (range 1-24 hours, mean 9 hours) according to their clinical conditions.

In each patient, an anteroposterior CXR in supine position was acquired with a portable x-ray equipment.

Twelve patients also had a chest CT scan performed within 48 hours before or after the admission. The indications for CT scan were : acute abdominal pain (5 patients) and evaluation of cancer pain (3 patients). Other four patients had a CT scan obtained for respiratory symptoms at outpatients centers before admission.

Two radiologists evaluated de-identified CXRs. The first radiologist with 35-year experience in practice and research on pulmonary imaging (M.G.) and the second with 15-year experience (S.M). The following features were assessed:

1)negative or positive for pneumonia.

The two readers were advised to consider as positive only cases in which they have a high level of confidence on the base of their experience.

In positive cases the following characteristics of the lesions were recorded

2)unilateral or bilateral lung involvement
3)topography 3a :upper or lower fields dividing each lung with a horizontal line passing through the center of the hila 3b : diffuse 3b: predominantly peripheral, non peripheral or mixed
4)density 4a : non consolidative ground-glass or inhomogeneous opacities 4b: condolidative nodular-type or triangular 4c: mixed consolidative e non-consolidative
5)presence of pleural effusion

The evaluation of point 1 (negative or positive) was carried out separately by the two readers and agreement was calculated.

The other points were evaluated in consensus.

The cases in which there was no agreement on point 1 were not further evaluated for the other points.

A new session was carried out to evaluate the quality of CXRs after a post processing combing fast and adaptive bidimensional empirical mode decomposition (FAMED) [12] and homomorphic filtering [13] with Contrast Limited Adaptive Histogram Equalization (CLAHE) [14]. Fig. 1 shows an example of CXR image post processed.

**Figure 1.**
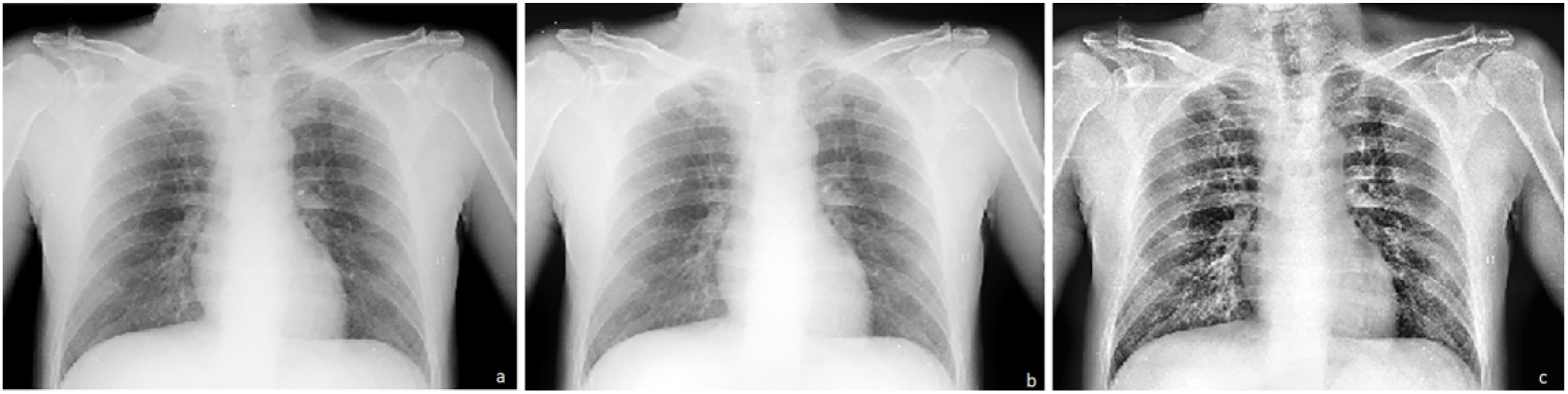
An example of x-ray image elaborated with the proposed approach (a) initial, (b) after the application of FABEMD and homomorphic filtering, (c) final image after the application of CLAHE algorithm.

Two radiologists both with 5-year experience (T.D. and G.C.) who were aware of the results of the baseline CXR analysis, performed this evaluation in consensus (readers 3 and 4).

Quality was assessed by comparing case by case both the native and the post-processed CXRs. The readers 3 and 4 were asked to evaluate the improvement of lesion detectability if present.

## STATISTICS

Agreement between the two readers regarding positive or negative CXR was evaluated and K-values were calculated.

## RESULTS

### POSITIVE vs NEGATIVE

Reader 1 recorded 50 cases out of 72 as positive (69%) and 22 (31%) as negative.

Reader 2 recorded 52 cases as positive (72%) and 20 as negative (28%).

Agreement was almost perfect with K 0.935, p < 0.001.

### CHEST X-RAY FINDINGS

CXR findings were evaluated by consensus in 50 cases considered positive by both readers.

- Lung involvement was unilateral in 25 out of 50 cases (50%), and bilateral in 25 (50%) (fig.2).
- Regarding topography, lesions were located exclusively in lower lung in 27 patients (54%), and both in upper and/ or lower lung in 23 (46%) (fig. 3).
- A prevalent peripheral distribution of lung abnormalities could be seen in 24 patients (48%). Lesions were evaluated as non-peripheral location in 11 (22%), and with mixed pattern (both peripheral and non-peripheral) in 15 (30%).
- Regarding density, non-consolidative pattern was present in 22 cases (44%) (Fig.4), consolidative in 9 (18%), and mixed in 19 (38%)(fig. 4). Single or multiple nodules could be seen as the only or prevalent pattern in 7 patients (14%). Triangular lesions were detected in 3 cases (6%).
- Pleural effusion was recorded in 2 patients (4%).

**Fig. 2.**
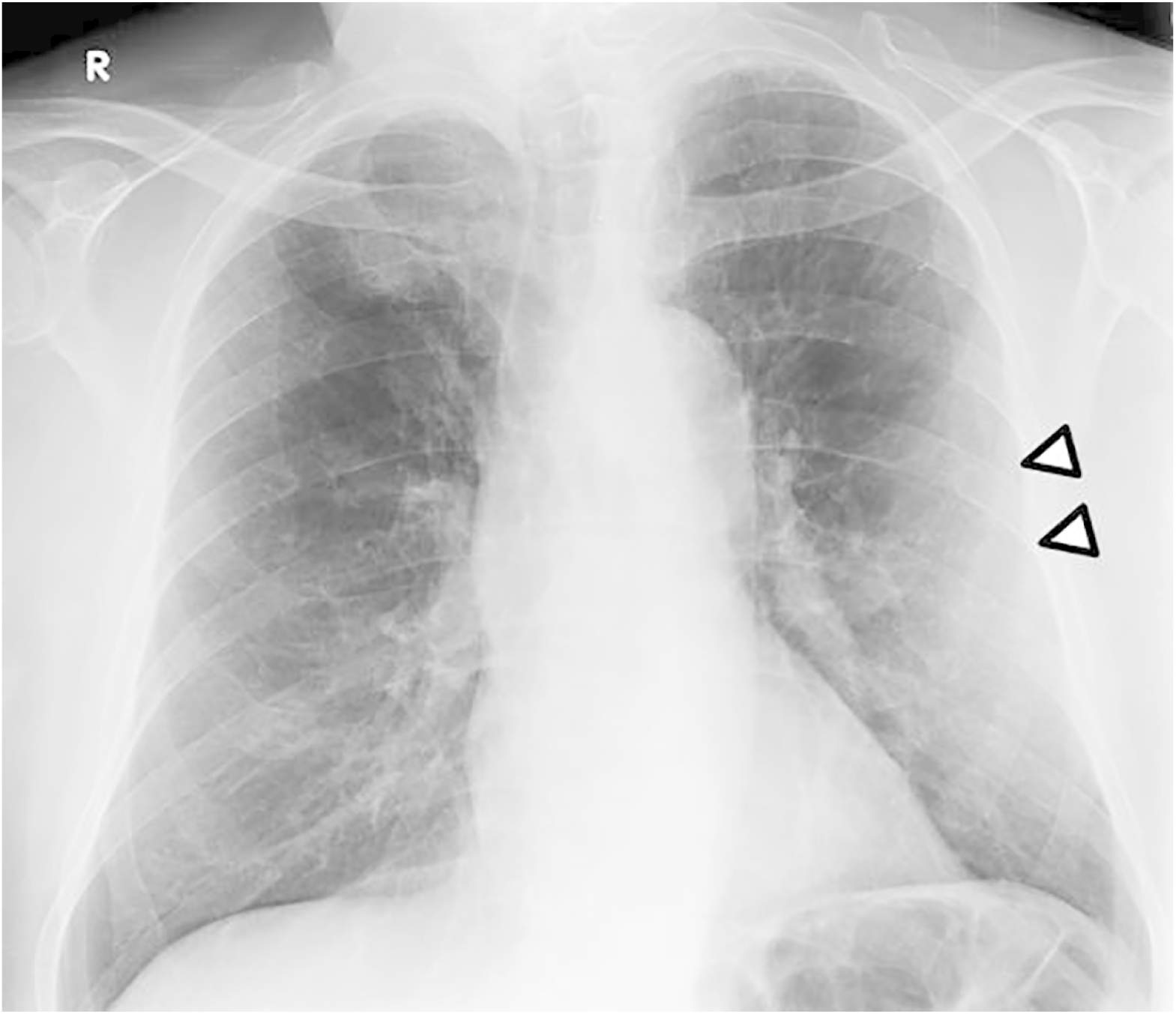
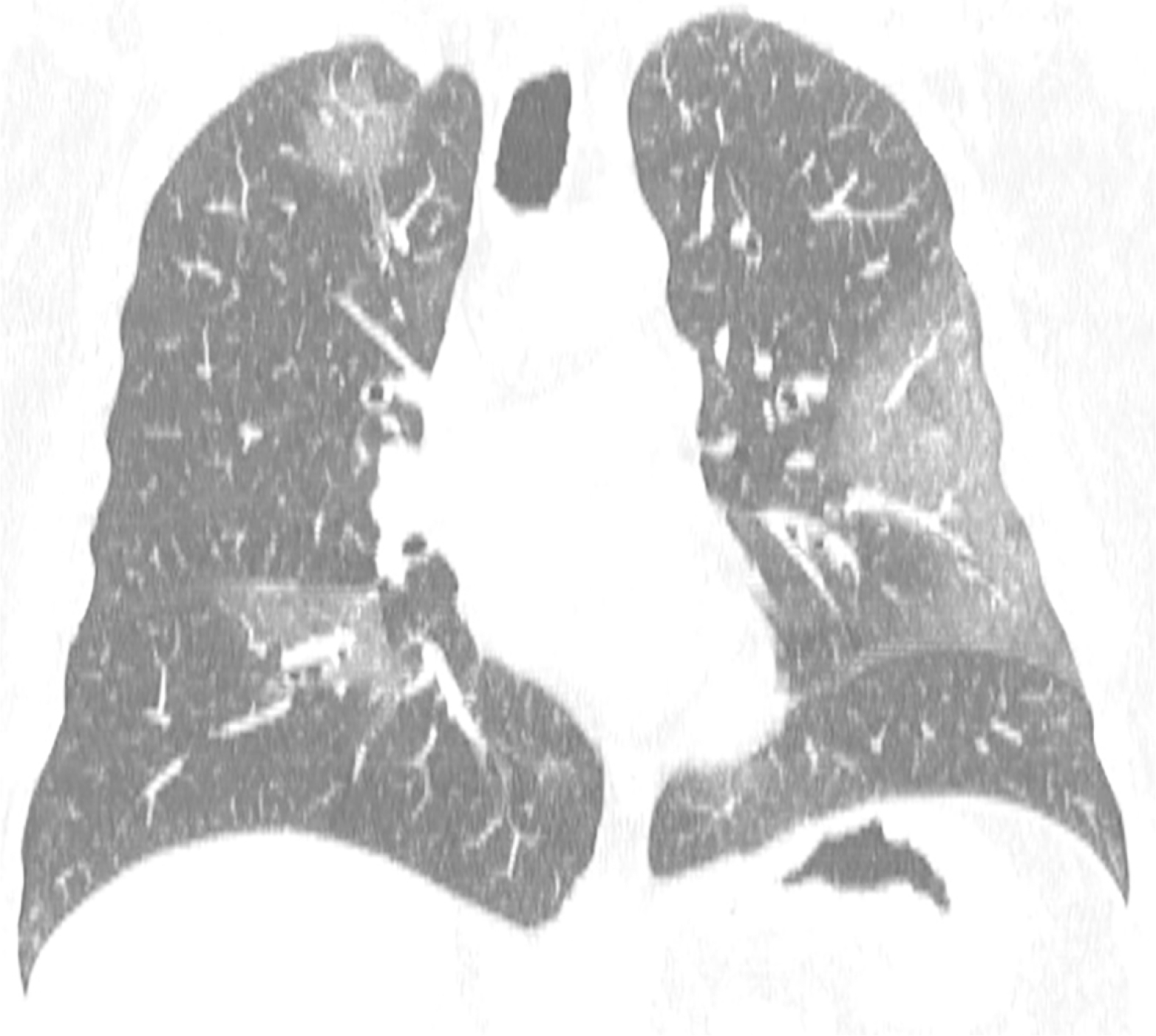
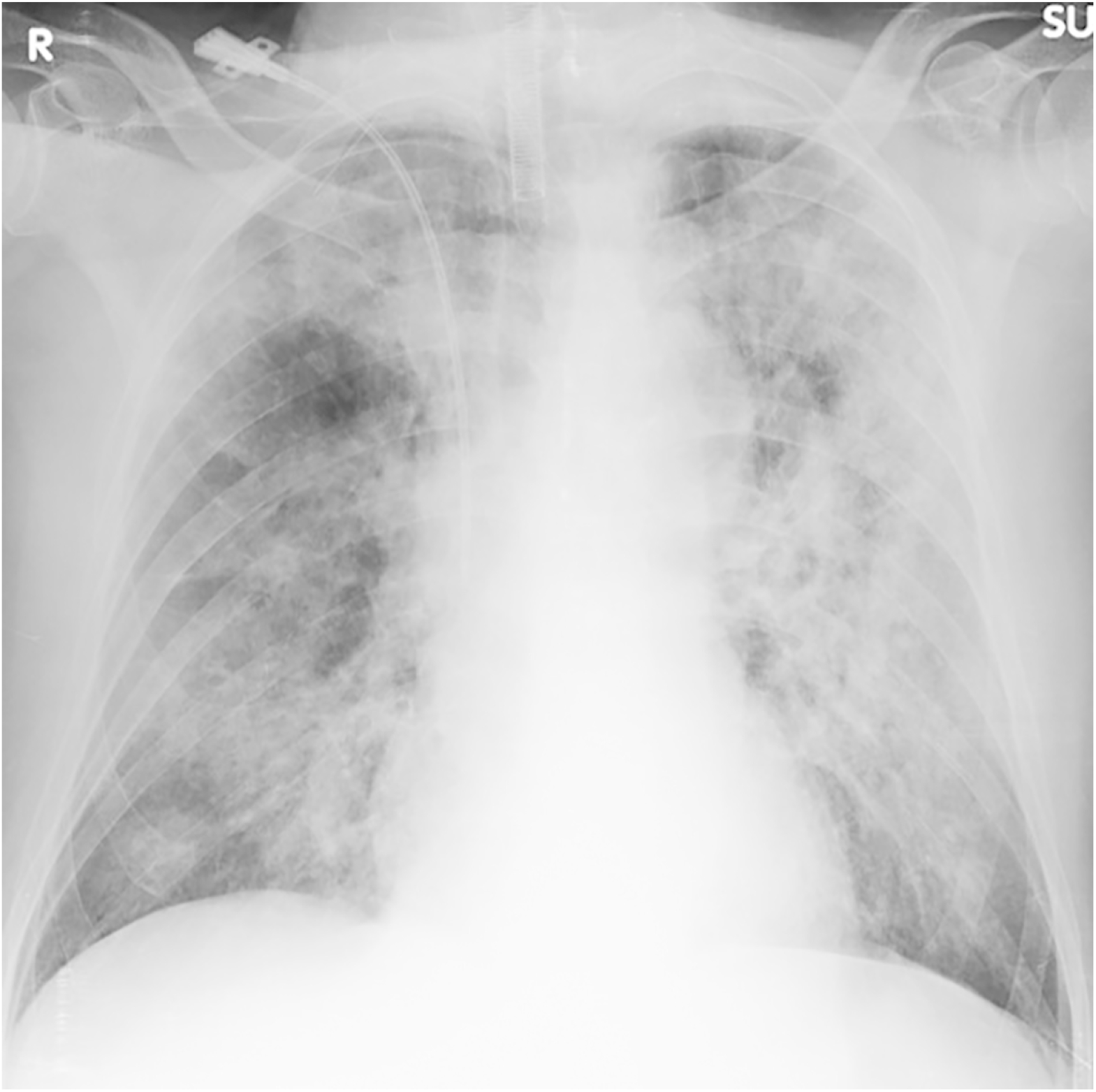
(a) In a 78 year-old male patient with fever and slight dyspnea chest x-ray shows a large non-consolidative lesion in the peripheral left lung (arrowheads). (b) Coronal reconstruction of CT-scan of the same patient performed 3 hours after, depicted small ground glass lesions in the right lung, not clearly visible on chest x-ray. (c) 82 year-old male patient with diffuse and bilateral lung involvement.

**Fig. 3.**
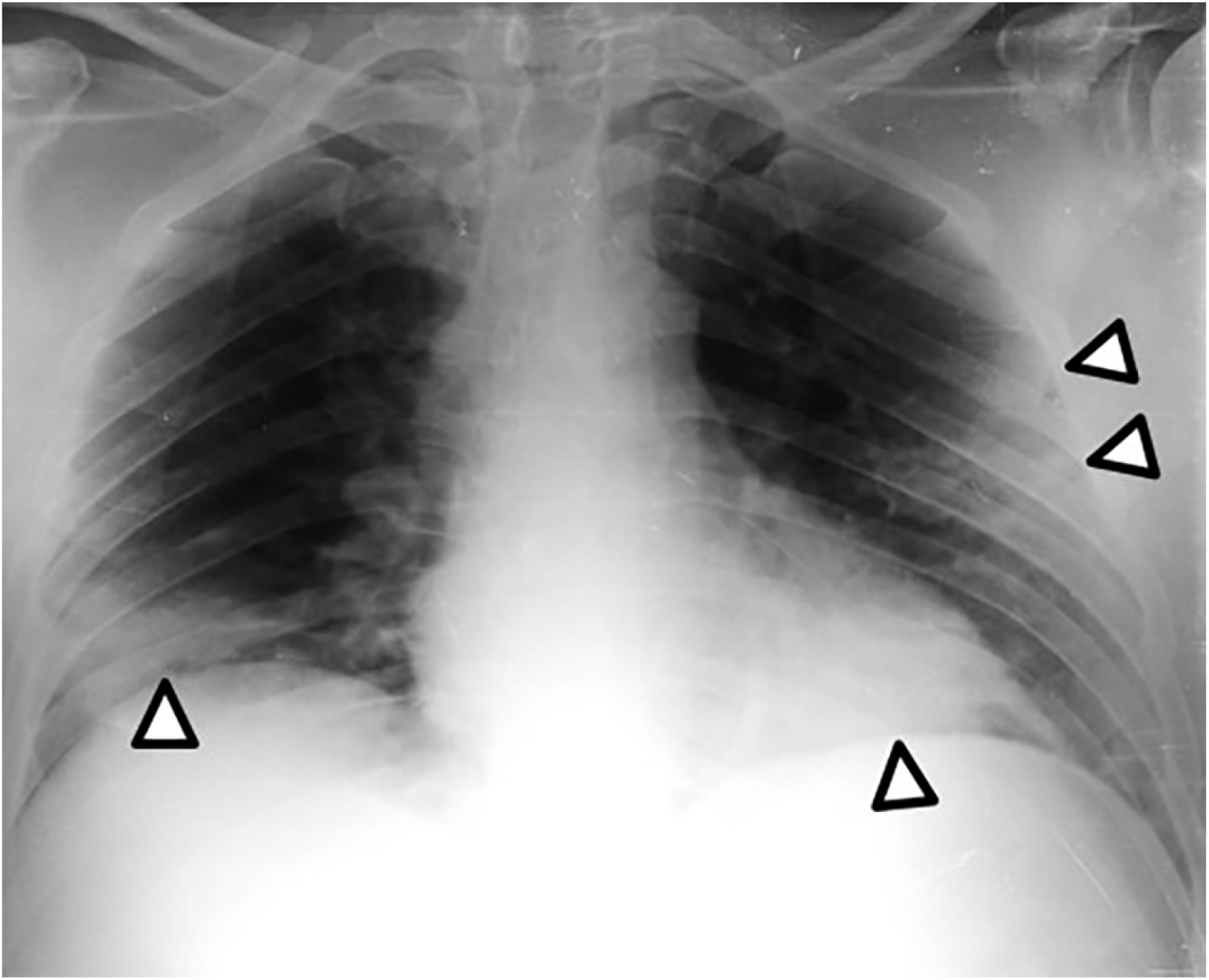
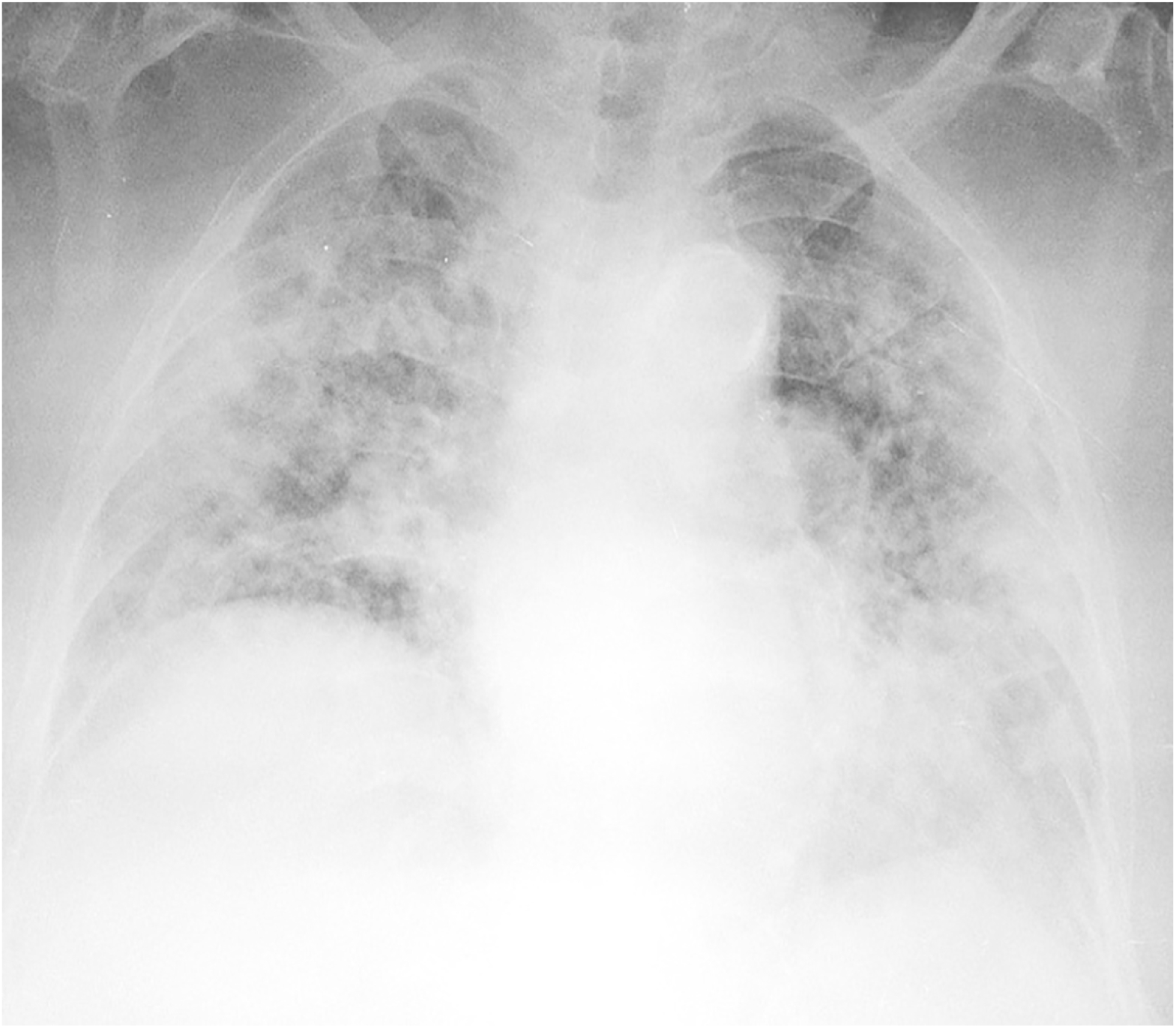
(a) 49 year-old male patient with bilateral lung involvement. In both lower lungs, the lesions are consolidative, nodular and triangular-shaped (arrowheads). (b) 92 year-old female patient with bilateral and diffuse involvement of upper and lower lungs.

**Fig. 4.**
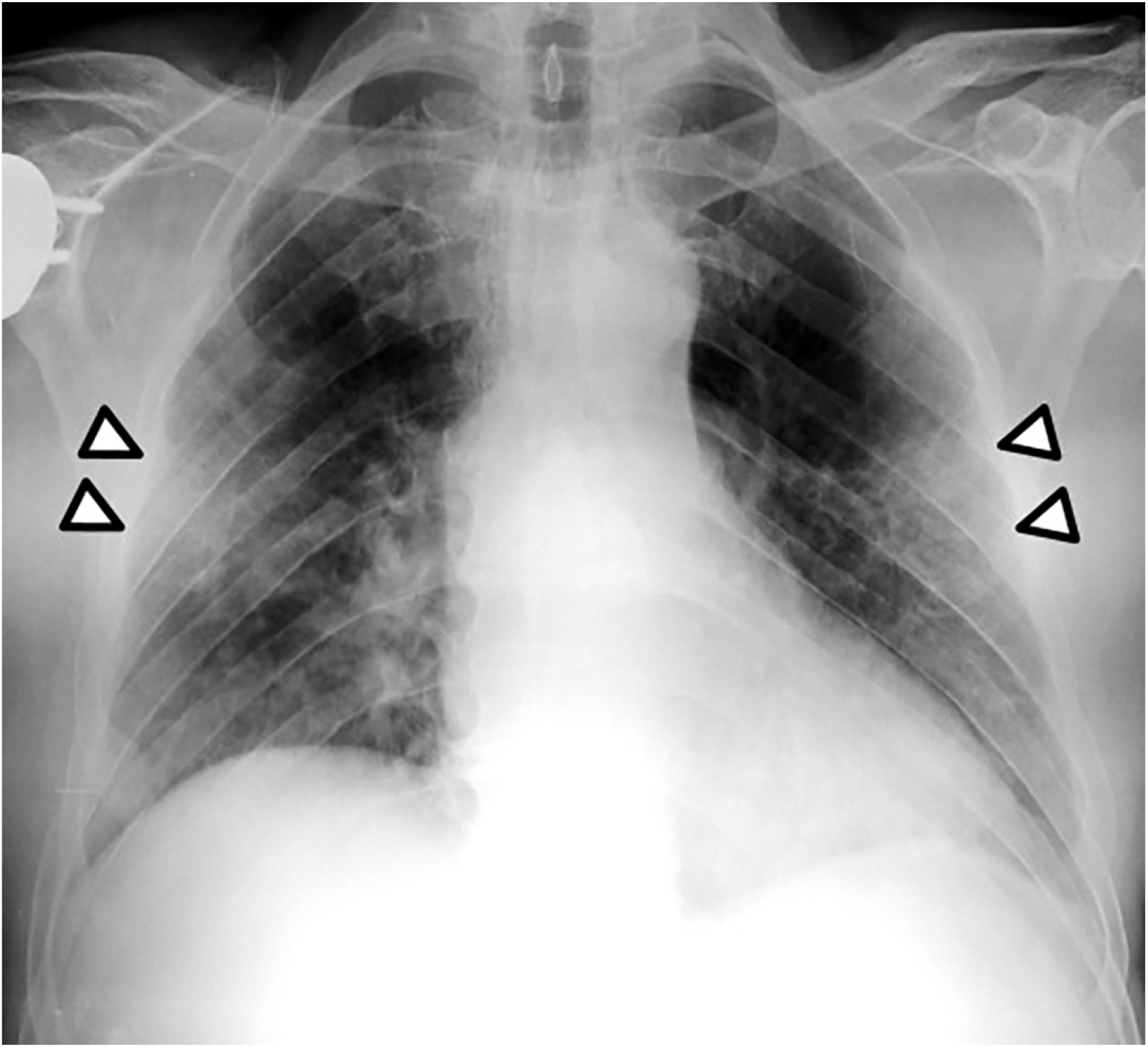
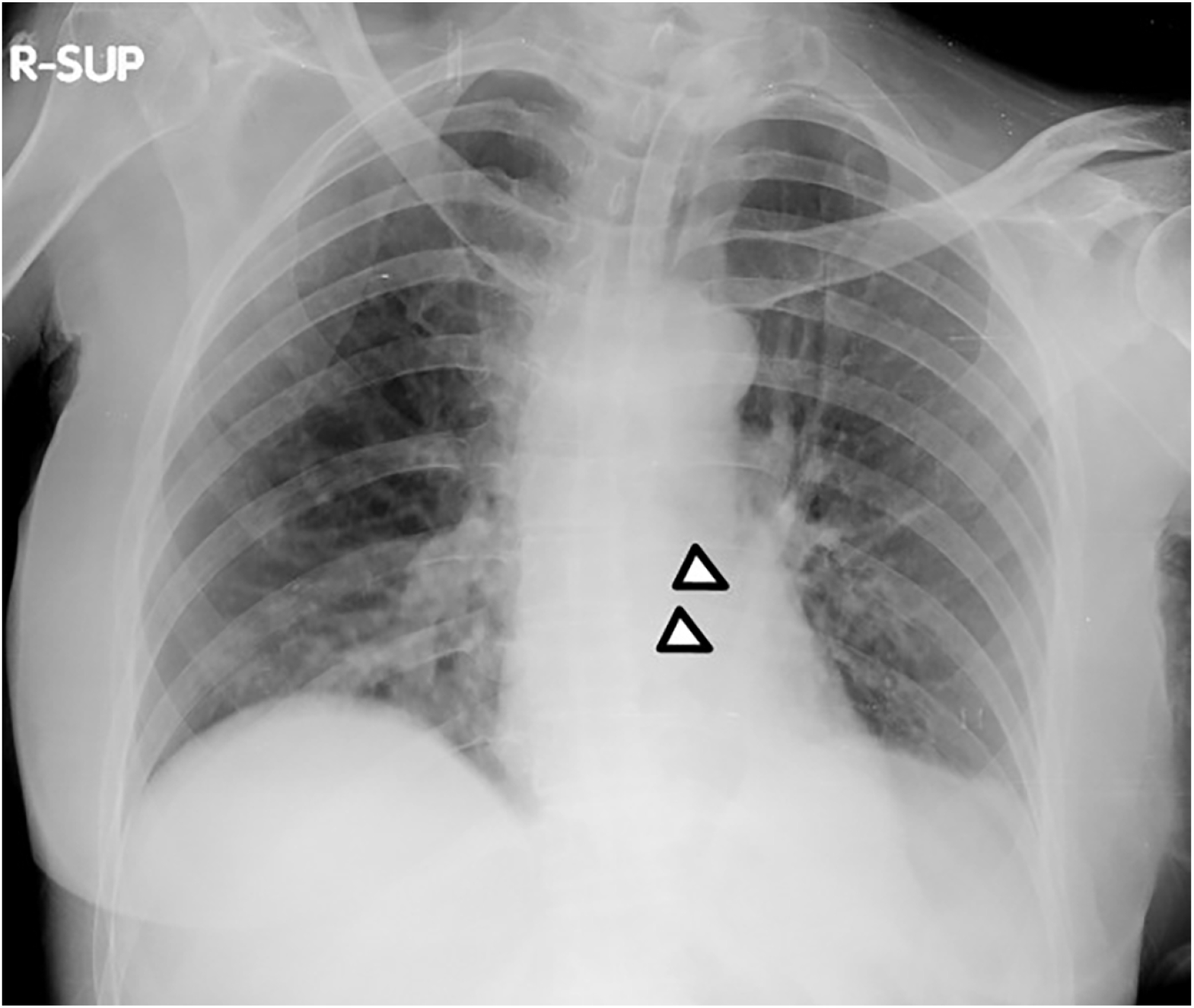
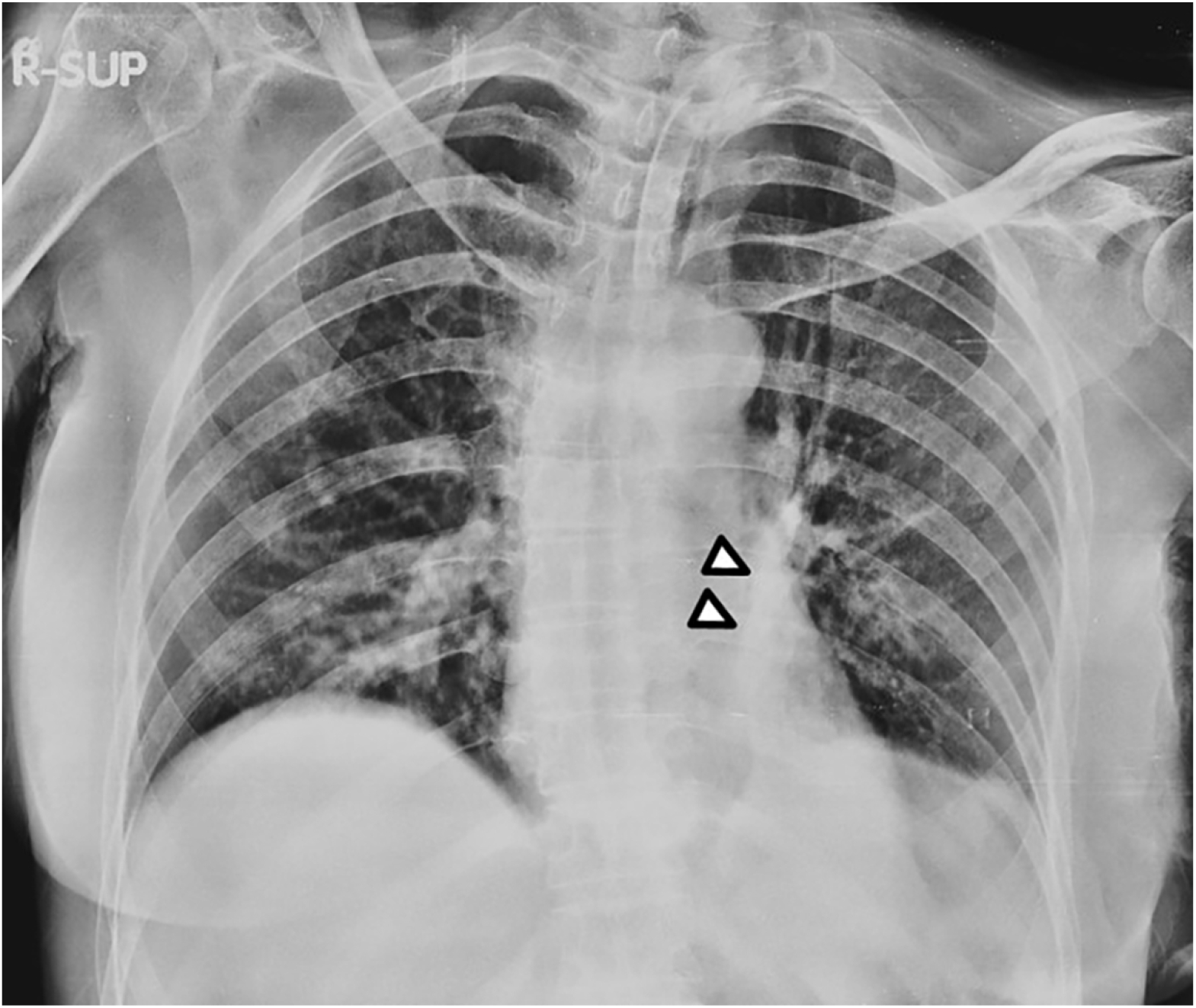
(a) 67 year-old male patient with bilateral and peripheral ground glass lesions (arrowheads). (b) 55 year-old female patient with a left lower lobe consolidation (arrowheads). (c) This finding is better appreciable on the post-processed image (arrowheads).

## EVALUATION OF POST-PROCESSED CXRs

The two readers detected further lesions not visible on native CXR, in 6 cases evaluated as positive by readers 1 and 2, and one in a patient evaluated as negative by both readers 1 and 2. The two cases with absent agreement between readers 1 and 2 were recorded both as negative by readers 3 and 4.

## DISCUSSION

Currently, the imaging literature of COVID-19 lung involvement is dominated by CT [2-9]. On the other hand, we found only a recent paper by Wong et al [11]. describing CXR features in a cohort of 64 patients.

Although CT owns a sensitivity higher than 90% in detecting Covid-19 pneumonia also in asymptomatic or slightly symptomatic patients, it is not without important drawbacks.

First, CT is not specific and diagnosis of COVID cannot be made by CT but needs of RT-PCR positivity according to WHO [1]. In addition COVID patients with negative CT exist [1-9].

Second, the use of CT requires a long and difficult sterilization of CT room and patient track to CT room, increasing the risk of COVID diffusion among operators and other high-risk non-COVID-19 patients (e.g. immune-incompetent or with cancer) that needs of not deferrable CT examination (e.g. restaging of cancer).

Third, the cost of CT and the limited availability of CT scanners or logistic problems could not allow the extensive use of this imaging technique. In particular, this monitoring activity could become very critical at the peak of new cases in any hospital. For those reasons, since the beginning of pandemic in our province (Messina-Italy) we started a protocol based on CXR at bed of patients admitted in our hospital.

In our series, we have found a sensitivity of CXR between 69 and 72% which is good enough to state that this low-cost, low-risk technique should restrict CT as problem solving examination only. Our data support advises of North-American Radiological Society Panel Expert to avoid CT in every suspect of COVID19 patients to reduce risks and costs without a clear advantage for patients [10].

Our sensitivity is high for CXR, but such a result can be explained by considering that our admission criteria did not include patients with no or mild symptoms.

Nevertheless our sensitivity is very similar to the one reported very recently by Wong et al. in a cohort of 64 patients [11].

Not surprisingly CXR findings represent the counterpart of CT features of COVID-19 lung [2-9, 15].

However in our cohort bilateral and prevalent peripheral distribution of abnormalities is not so striking as in CT studies.

Probably the high rate of unilateral involvement could be correlated to the lack of sensitivity for small ground-glass opacities visible only on CT (Fig. 2a, b).

In 10 out of 50 patients nodular, multi-nodular and triangular lesions could be seen (Fig. 3a, 4b-c). This type of lesions has been already described as atypical on CT scan [15].

This work has some limitations.

First, our cohort, due to admission criteria, does not include asymptomatic or mild patients where CT scan can detect ultra-early and early findings of pneumonia [16], and then we cannot state which is the sensitivity of CXR in these patients.

However, a recent research letter reported that CXR film can detect pneumonia also in some patients with mild COVID-19 disease [17]. Further studies are necessary on this direction.

Second, we cannot report the outcome of patients because at the time of writing many of them have not concluded the disease cycle.

Third, we have not obtained correlation with the known standard of reference for COVID-19 imaging which is chest CT since only a small sub-sets of our patients underwent CT. No attempt to correlate CXR and CT findings of this small sub-set of patients was made but some anecdotal cases have been used in our representative figures. However the almost perfect agreement between our two independent readers support the reliability of our results also considering similar results published by Wong et al. [11].

Quality of chest x-ray films obtained at bed in supine position cannot be perfect in a significant number of cases, particularly in old patients suffering of dyspnea. This limitation drove the development and clinical test of a post processing tool to improve the contrast resolution of CXR. Quality evaluation of post-processed CXRs was made either on the base of rigorous metrics (entropy) either on subjective evaluation. Both types of analyses showed a significant improvement of quality. Interestingly, in 7 cases out of 72 patients (≅10%) further lesions not detected on baseline CXR could be seen on post-processed images (Fig.5).

**Fig. 5.**
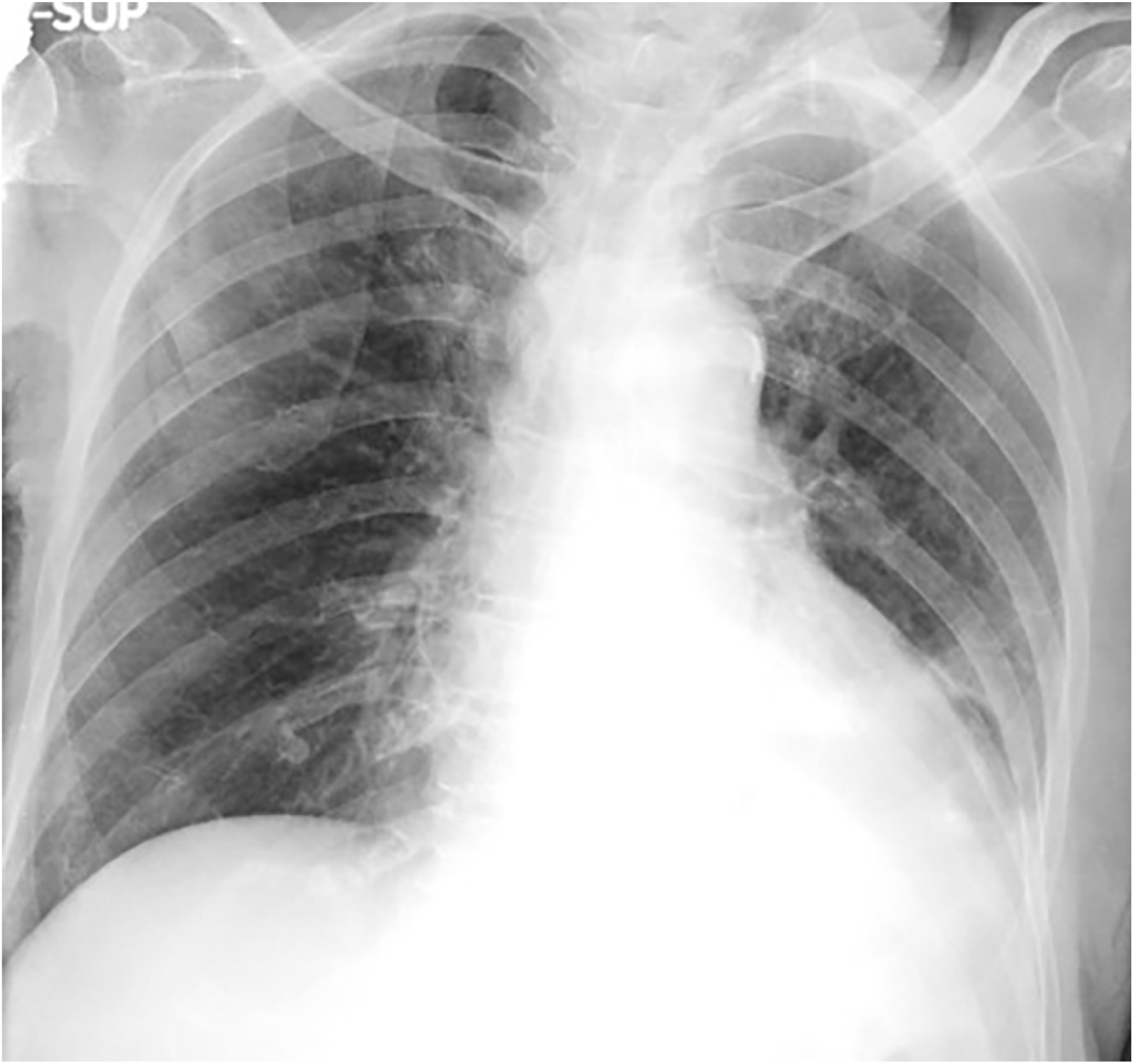
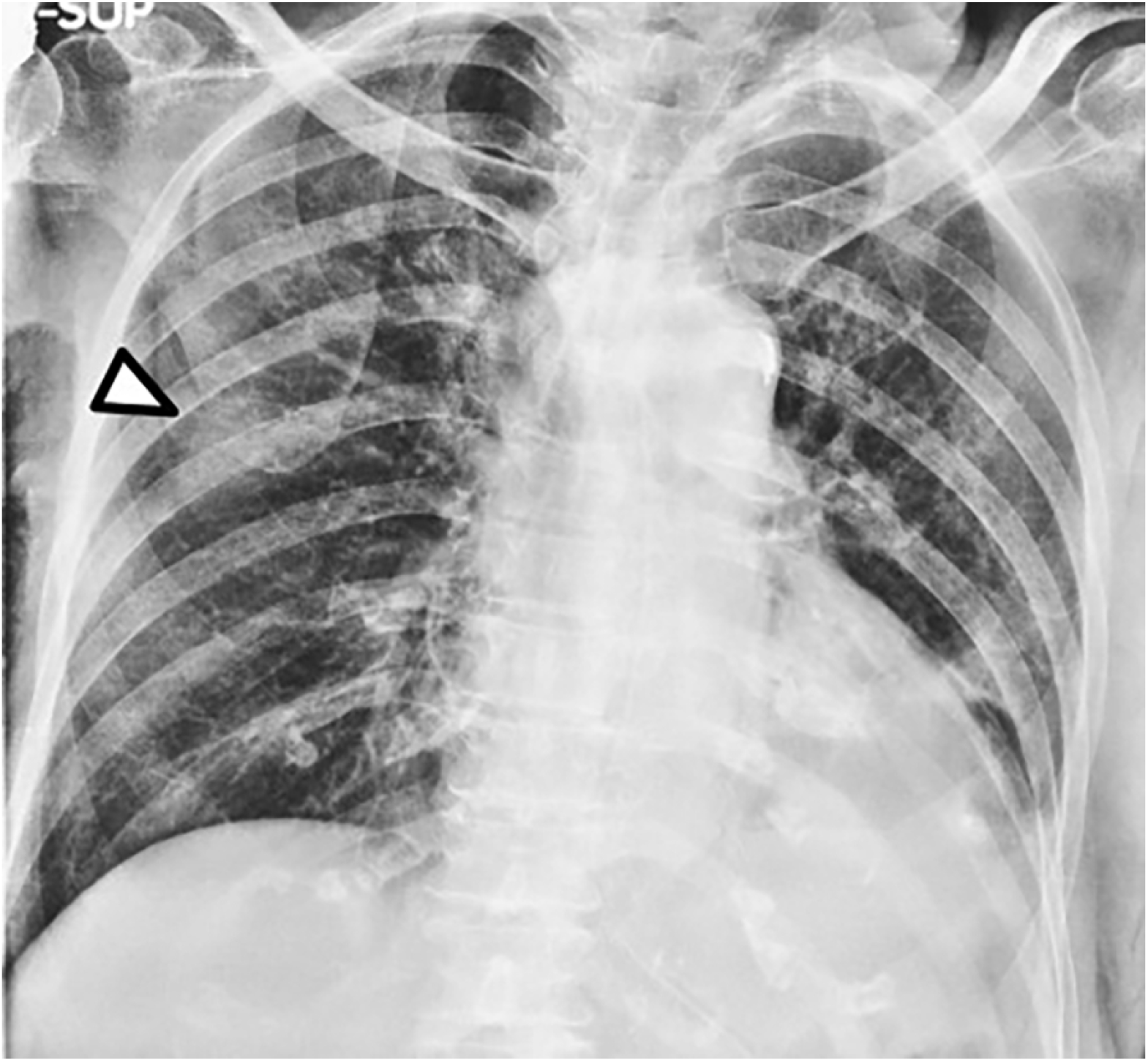
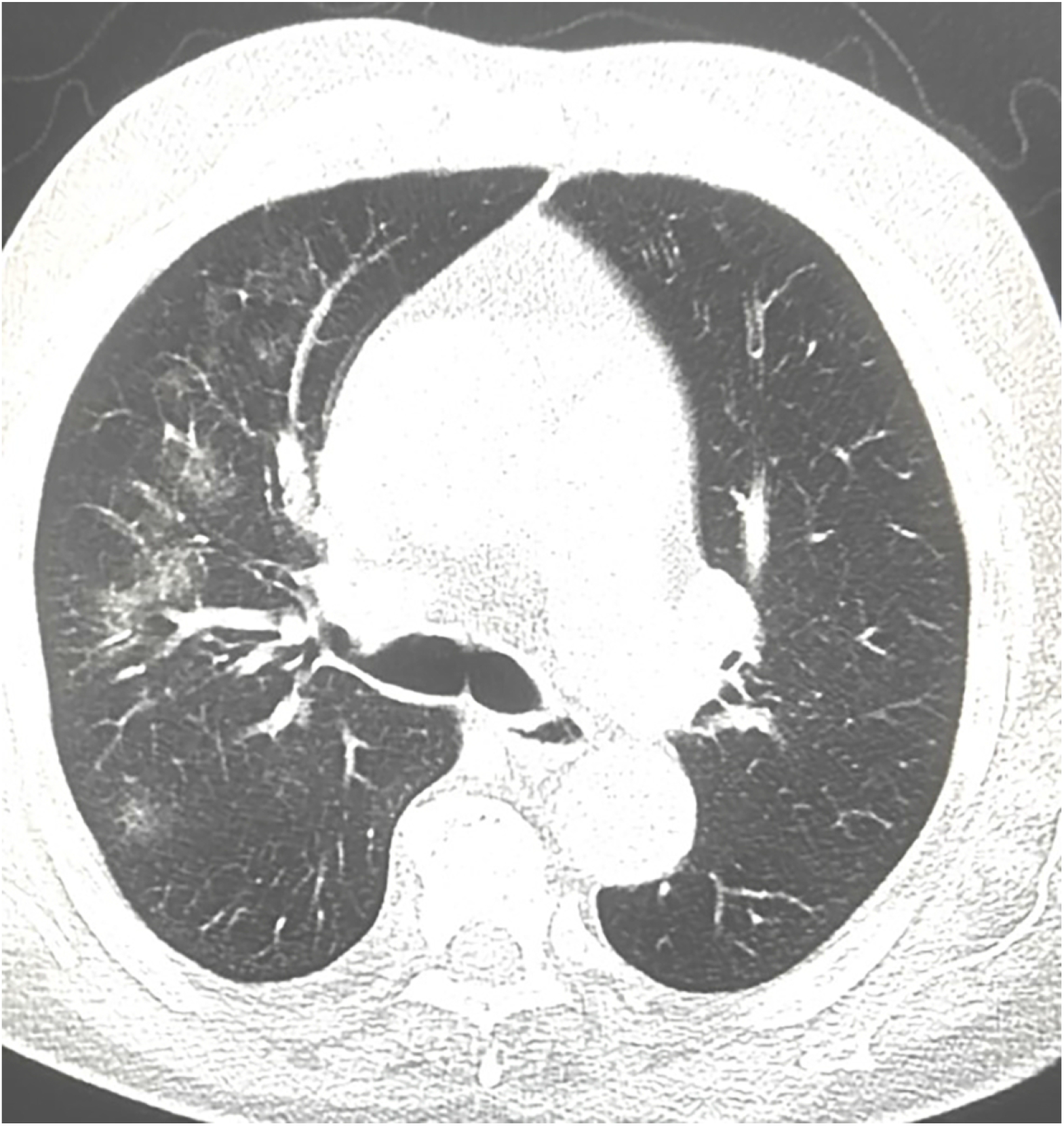
87 year-old male patient with a small left pleural effusion. (a) On baseline CXR, right lung was consirate negative for parenchimal lesions by both readers. (b) Post-processed image revealed a ground-glass opacity in the right lung (arrowhead). (c) CT-scan, performed 6 hours later for upper abdominal pain, was extended to the thorax, which showed sparse ground glass opacities within upper lobe of the right lung.

In conclusion, our data show some novel characteristics of CXR findings in the most extensively group of COVID-19 patients who have to be studied during hospitalization, namely patients with not early disease:

1. CXR at bed owns a good sensitivity in detecting COVID-19 lung involvement in patients with not early disease.
2. Our data support the (RSNA) Panel Expert recommendations on imaging approach to COVID-19 pandemic based on CXR rather than protocols based on extensive use of CT [18].
3. The use of proper post-processing tools can improve quality of CXR and detection of lesions.

## Data Availability

Data available on request to the corresponding author.

